# Data-driven discovery of a clinical route for severity detection of COVID-19 pediatric cases

**DOI:** 10.1101/2020.03.09.20032219

**Authors:** Hui Yu, Jianbo Shao, Yuqi Guo, Yun Xiang, Chuan Sun, Ye Yuan

## Abstract

The outbreak of COVID-19 epidemic has caused worldwide health concerns since Nov., 2019. A previous study described the demographic, epidemiologic, and clinical features for infected infants. However, compared with adult cases, little attention has been paid to the infected pediatric cases. Severity detection is challenging for children since most of children patients have mild symptoms no matter they are moderately or critically ill therein.

## Introduction

The outbreak of Coronavirus Disease 2019 (COVID-19) epidemic has caused worldwide health concerns since Nov., 2019 [1]. A previous study [1] described the demographic, epidemiologic, and clinical features for infected infants. However, compared with adult cases, little attention has been paid to the infected pediatric cases [2-6].

Severity detection is challenging for children since most of children patients have mild symptoms [2] no matter they are moderately or critically ill therein.

## Methods

For this retrospective study, we identified 105 infected children admitted to Wuhan Children’s Hospital, the sole designated hospital in Wuhan for COVID-19 children patients, from Feb. 1 to Mar. 3, 2020. The epidemiological, clinical laboratory, and outcome data were extracted from the medical records of these patients. The study was approved by Wuhan Children’s Hospital Ethics Committee.

Throat-swab, anal-swab or urine specimens were taken at admission for real-time RT-PCR, which was performed by Wuhan Huada Gene Biology. Symptoms on admission were collected together with laboratory results, chest radiography and CT findings, treatment received for COVID-19 and clinical outcomes.

Severe patients are generally manually detected according to the guideline from National Health Commission, China, using clinical symptoms including shortness of breath, assisted respiration, apnea, cyanosis, dehydration, and progressive increase of lactate, etc.

In this study, a supervised decision-tree classifier was developed. All clinical measurements from the last available date were used as features and set ‘mild’ and ‘severe’ as labels. “-1” was used to complement the incomplete clinical measures to avoid bias. We used standard F1-score [4] to evaluate the performance of the classifier. We started from one feature and increased the number of features in the clinical route, until the F1-score converges. We picked the classifier with less incomplete measurements for all the patients, when two classifiers with the same number of features performed the same.

## Results

105 children aged 1-16, including 64 males and 41 females, were infected with COVID- 19, of which 8 were critically ill. The youngest was 1 day after birth, and the oldest was 15-year-old. Male infection rate (60.95%) is higher than that of the female (39.05%). This is opposite to a previous report [2]. Children over 6-year-old have the highest infection rate (60.95%). All the children lived in Wuhan.

Based on the current available clinical data, we discovered a clinical route that can achieve 100% F1 score (shown in Figure 1A). Figure 1B depicts the mild and severe children patients over the proposed two-feature based clinical route. As a result, we have extracted merely two features, i.e., Direct Bilirubin (DBIL) and alaninetransaminase (ALT), by which 8 critically ill pediatric cases can be precisely identified from other 97 mild patients.

**Figure 1.**
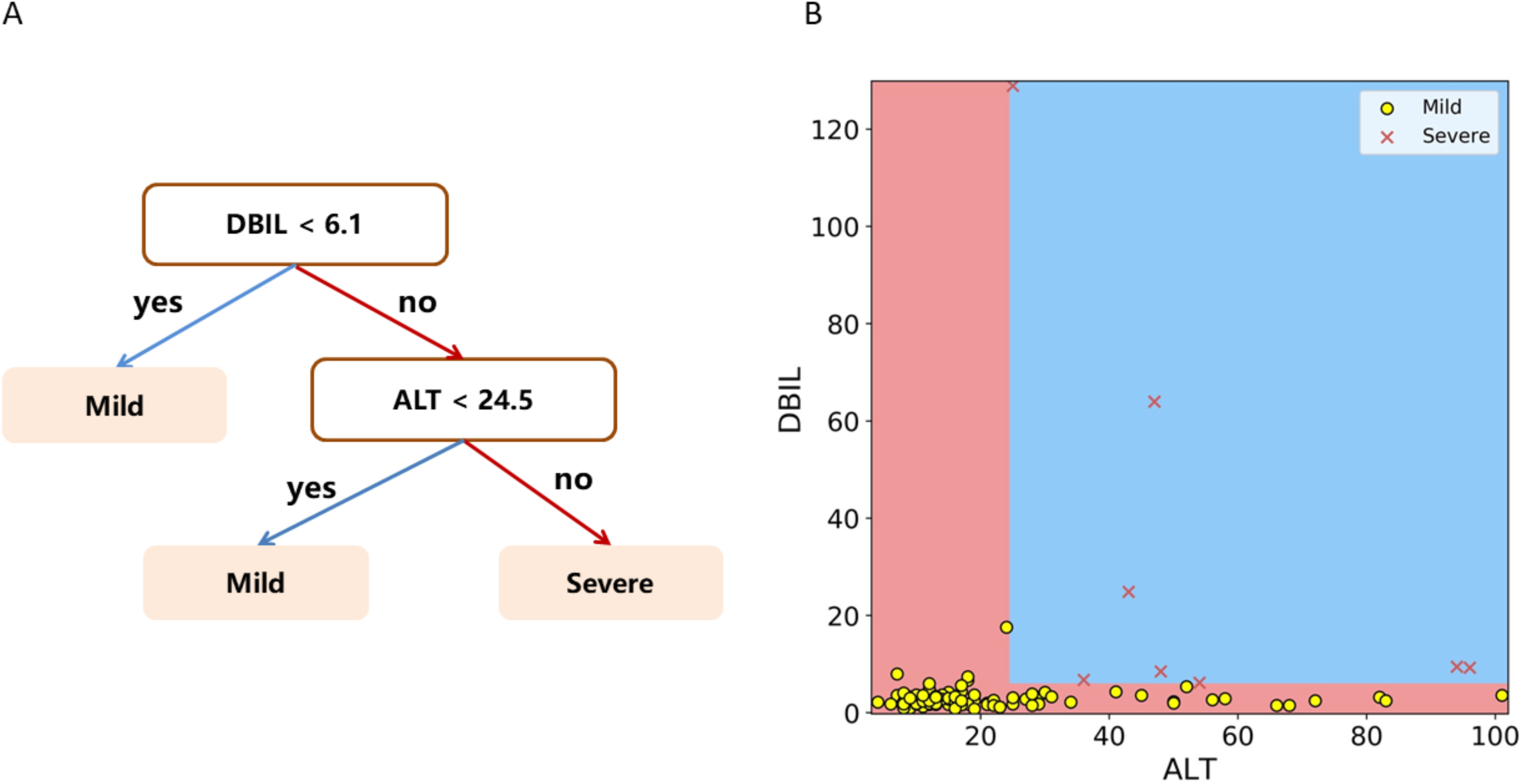
A. A diagram showing identification of severe cases using the discovered clinical route. These two features are automatically selected from over 300 features from the proposed classification algorithm; B. Visualization of mild and severe patients over the selected features.

## Discussion

Due to the scantiness of clinical data from confirmed COVID-19 children cases, especially for severe ones, it is an urgent yet challenging mission to promptly distinguish the severe ones from the mild cases for early diagnosis and intervention. To this end, with the assistance of machine learning methods, we identified that DBIL and ALT, surfacing from over 300 clinical features, were able to serve as a combination index to screen out all the critically ill cases. Although the increase of DBIL and ALT has been reported to reflect tissue destruction or injury, for the first time, their combination is revealed as a precise indicator for the severity of COVID-19 pediatric cases, which is quite different from the discovered clinical route for adult [4].

The study was limited to a small number of patients from a single center in Wuhan. Further studies from multiple centers in a larger cohort would be beneficial to the validation of the proposed route as well as understanding of the disease.

## Data Availability

None

## Abbreviations

COVID-19: Coronavirus Disease 2019
DBIL: Direct Bilirubin
ALT: Alaninetransaminase

## Footnotes

DBIL: Direct Bilirubin; ALT: Alaninetransaminase

---

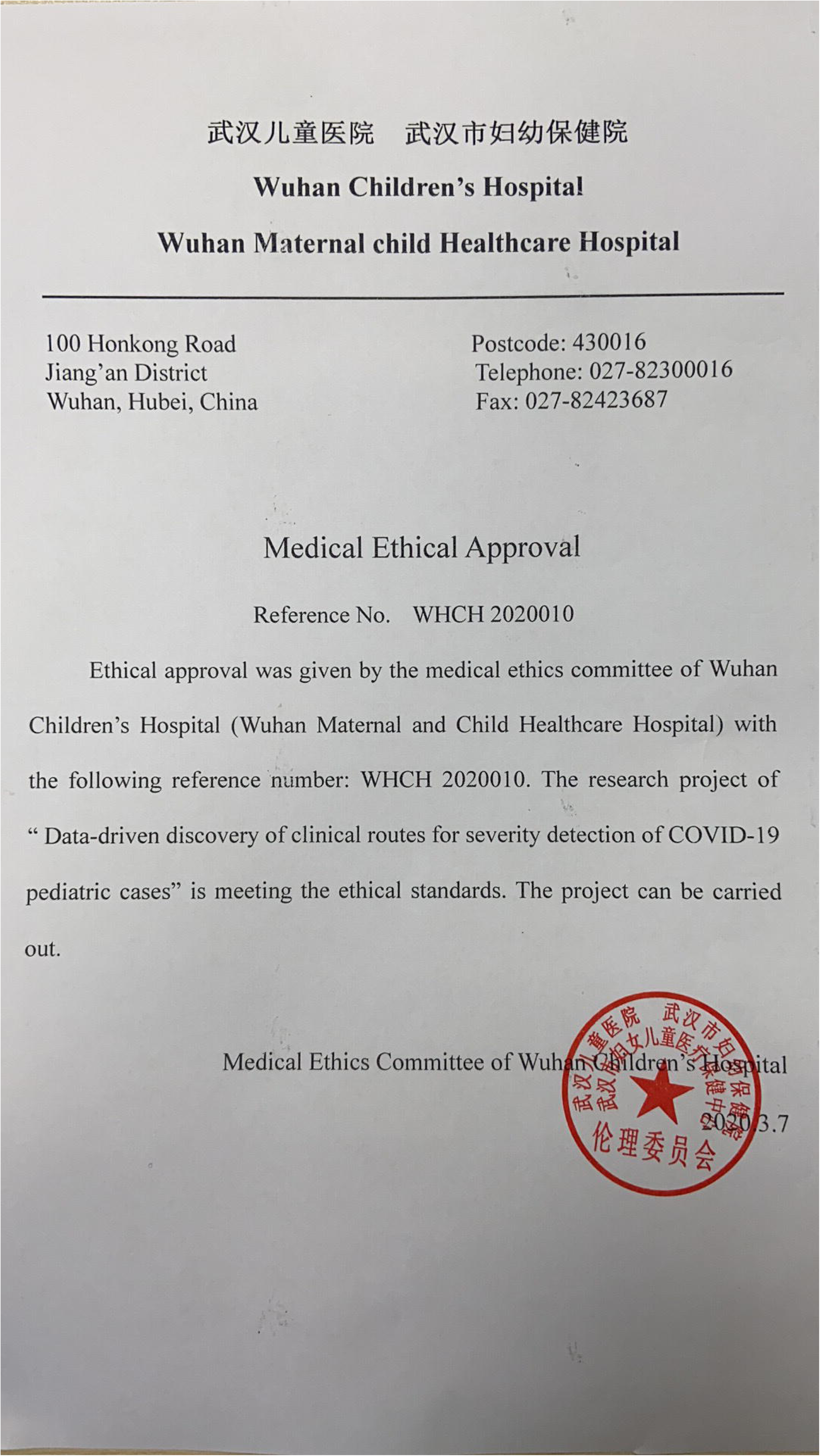

